# A longitudinal study of anxiety and depression in Belgium during and after the COVID-19 pandemic

**DOI:** 10.64898/2026.02.24.26347039

**Authors:** Thanh Thuy Bui, Stefaan Demarest, Camille Duveau, Lize Hermans, Guido Van Hal

## Abstract

**Background:** The COVID-19 pandemic, coupled with concurrent social instabilities, has raised concerns about the long-term impact on the population mental health. While existing studies have primarily focused on the acute phase, less is known about how anxiety and depression symptoms have evolved throughout prolonged societal disruption. This study aimed to identify distinct anxiety and depression symptom trajectories and to determine the individual, relational, and societal protective and risk factors that influence anxiety and depression scores among Belgian adults from 2020 to 2024.

**Methods:** We used longitudinal data from five waves of the COVID-19 Health Surveys and the BELHEALTH study (n = 10,063) among Belgian adults, collected between April 2020 and June 2024. Anxiety and depression were assessed using the Generalized Anxiety Disorder-7 and the Patient Health Questionnaire-9, respectively. Covariates were selected based on the social-ecological framework and included both time-invariant and time-dependent variables. Latent class linear mixed models identified subgroups with distinct trajectories. Multilevel linear mixed effects models examined associations between symptom severity and predictors across individual, relationship, and societal levels. The final model, selected based on the lowest AIC (Akaike Information Criterion), included the full set of covariates.

**Results:** Four depression and five anxiety trajectories were identified. While most participants maintained stable mild symptoms, 11.3% experienced increasing depression and 8.4% showed increasing anxiety over time. Financial difficulty, female gender, and younger age were overrepresented in moderate and severe symptom trajectories. Protective factors such as social support, satisfying social contact, and life satisfaction were associated with lower symptom severity. Over time, life satisfaction demonstrated an increasing protective effect, while the influence of social contact on reducing symptoms weakened progressively. Risk factors included financial and job-loss worry, loneliness, psychotropic medication use, and high mental health stigma.

**Conclusions:** Our results demonstrate persistent heterogeneity in mental health responses, with a substantial share of the population experiencing worsening symptoms years after the pandemic began. Public mental health strategies must therefore go beyond short-term crisis response, address long-term risks such as financial insecurity, social isolation, and stigma, while fostering individual and collective resilience.

## BACKGROUND

Mental health disorders, particularly anxiety and depression, have long been recognized as major contributors to the global burden of disease, imposing significant challenges on individuals, healthcare systems, and societies worldwide. The advent of the COVID-19 pandemic further exacerbated the already significant global burden of mental health disorders. In response to the unprecedented global health emergency, the World Health Organization (WHO) reported a staggering 25% increase in the global prevalence of anxiety and depression (1, 2). This sharp rise underscored the profound psychological toll of the pandemic, which was characterized by widespread fear of infection, loss of life, economic disruption, and drastic changes to daily routines and social interactions. In Belgium, a similar trend was observed; data from the COVID-19 Health Surveys indicated that anxiety symptoms among adults surged to 21% and depression symptoms rose to 17% during the initial lockdown period in 2020 (3). This figure was notably higher than the pre-pandemic levels recorded in 2018 (11.2% anxiety and 9.4% depression), highlighting the acute impact of the crisis on the nation’s mental well-being (4). Mental health trends associated with the pandemic’s progression and restrictive measures, at restrictions eased, some symptoms decreased to near pre-COVID levels due to improved coping, though certain subgroups remained significantly affected.

While there is extensive literature on the immediate psychological effects of the pandemic, including heightened stress, uncertainty, and social isolation, there remains a critical gap in understanding the long-term mental health consequences as society transitions to a post-pandemic phase (5, 6). Additionally, previous studies often capture only overall trends over time, leading to an oversight of the hidden heterogeneity in mental health patterns.

The transition to a post-pandemic world does not signify an immediate cessation of mental health challenges. Numerous factors continue to exert considerable psychological pressure on many individuals. Ongoing economic instability, the persistent threat of job loss or difficulty in securing employment, and the lingering effects of prolonged social isolation contribute to a sustained psychological burden. Furthermore, individuals who contract severe forms of COVID-19, or those who experienced significant loss and trauma during the pandemic, might remain at an elevated risk of developing persistent mental health issues, including chronic anxiety and depression. Compounding these challenges, research has consistently shown that certain demographic groups bore a disproportionate mental health burden during the pandemic. Women, young adults, and individuals from lower socioeconomic backgrounds were among those most adversely affected. However, it remains largely unclear whether these vulnerable groups are recovering at the same pace as others, or if pre-existing disparities are being further entrenched in the post-pandemic landscape (3, 7, 8). Therefore, a comprehensive understanding of these long-term mental health outcomes, particularly among diverse demographic segments, is critical for the development and implementation of targeted and equitable public health interventions designed to foster robust recovery and enhance community resilience in the aftermath of the pandemic.

Among the most prevalent long-term mental health outcomes observed during and after the pandemic are symptoms of anxiety and depression (9, 10). To effectively address these complex and evolving challenges, it is crucial to meticulously examine how anxiety and depression symptoms evolve over extended periods and how various pre-existing protective and risk factors interact to shape these long-term mental health trajectories. The social-ecological model offers a valuable and comprehensive framework for this endeavor, conceptualizing mental health risks and protective factors as operating across multiple, interconnected levels: individual, relationship, and societal (11). Individual factors encompass demographic characteristics, personal health history, and psychological attributes. Relationship factors relate to social support networks and relationship quality, while societal factors include perspective about mental health, trust in authority and public policies. Recognizing that these multifaceted factors are dynamic and can fluctuate significantly over time, a longitudinal study design is indispensable for capturing the inherent complexity of post-pandemic mental health. A longitudinal approach allows for the systematic tracking of mental health indicators from the pandemic’s onset through the subsequent recovery phase. This enables a thorough and nuanced evaluation of the long-term effects of the COVID-19 crisis on anxiety and depression, providing insights into patterns of adaptation, vulnerability, and resilience.

Consequently, this study aims to address the identified knowledge gaps by pursuing two primary objectives. Firstly, this research seeks to describe the temporal evolution of anxiety and depression symptom scores over time, specifically examining how these trajectories differ across various demographic groups within the Belgian adult population. Secondly, this study endeavors to identify key protective and risk factors, situated within the individual, relationship, and societal domains of the social-ecological model, that significantly influence the course of anxiety and depression symptoms over the long term. By achieving these objectives, this research will contribute valuable evidence to inform the development of targeted public health strategies, clinical interventions, and policy initiatives aimed at mitigating the long-term mental health impact of the COVID-19 pandemic and strengthening societal preparedness for future public health emergencies.

## METHODS

### Study design, setting and participants

This investigation employed a secondary analysis of two comprehensive datasets collected by Sciensano, the Belgian institute for public health. The first dataset, the COVID-19 Health Surveys, comprises a series of cross-sectional surveys administered between April 2020 and March 2022. These surveys were specifically designed to monitor the evolving mental health impact of the COVID-19 pandemic on the Belgian population (12). The second dataset originates from the BELHEALTH project, a longitudinal follow-up study initiated in September 2022. The BELHEALTH project prospectively tracks participants from the earlier COVID-19 Health Surveys to assess their mental health status and trajectories in the post-pandemic period (13).

### Data collection and management

Data was collected at five critical time points, spanning the period from April 2020 to June 2024, to capture the dynamic nature of mental health during and after the pandemic. Table 1 provides a detailed overview of these data collection waves, including the specific timing, the prevailing pandemic-related situation, the source dataset, and the number of participants at each wave (14, 15). To ensure data consistency and enable longitudinal tracking, participants across these two data sources were meticulously matched using unique identification numbers. A key inclusion criterion for the analytical sample was the completion of at least two survey waves, resulting in a final sample of 10,063 participants, a prerequisite for reliably estimating time-varying outcomes and modeling individual trajectories of mental health symptoms.

**Table 1.** Data collected at five key time points from 2020 to 2024.

|  | **Time** | **Situation** | **Data sources** | **N** |
| --- | --- | --- | --- | --- |
| T1 | April 2020 | Initial COVID-19 pandemic | COVID-19 Health Survey | 49,215 |
| T2 | March 2021 | Mid pandemic | COVID-19 Health Survey | 20,351 |
| T3 | June 2022 | End of pandemic restrictions | COVID-19 Health Survey | 16,916 |
| T4 | February 2023 | Post-pandemic | BELHEALTH cohort project | 8,377 |
| T5 | June 2024 | Post-pandemic | BELHEALTH cohort project | 6,631 |

### Measurements

#### Outcome variables

The primary outcomes of interest in this study were symptoms of anxiety and depression. Anxiety symptoms were assessed using the Generalized Anxiety Disorder 7-item scale (GAD-7), a widely validated self-report measure. Each of the 7 items is rated on a 4-point Likert scale, ranging from 0 (not at all) to 3 (nearly every day), based on the respondent’s experiences over the preceding two weeks. Total GAD-7 scores range from 0 to 21, with higher scores indicating greater anxiety severity (16).

Depression symptoms were measured using the Patient Health Questionnaire 9-item scale (PHQ-9), another well-established self-report instrument. The PHQ-9 consists of 9 items rated on a 4-point Likert scale (0–3) reflecting symptom frequency over the past two weeks. Total PHQ-9 scores range from 0 to 27, with higher scores signifying more severe depressive symptomatology (17).

#### Covariates

A comprehensive set of covariates was selected to identify potential protective and risk factors influencing anxiety and depression symptoms over time. These predictor variables were chosen based on the established social-ecological framework, which categorizes influencing factors into multiple domains. The selected covariates were grouped as follows:

**Domain 1: Individual Factors**: This domain included 3 sub-groups

1. Demographic variables such as age (categorized as 18-29, 30 – 49, 50 – 64, 65+ and also treated as continuous), gender (male, female), highest level of education attained (low educated at most secondary school diploma, high education), household type (living alone; as a couple without children; as a couple with children; living alone with children; with my parent, family, friends or acquaintances), perceived income sufficiency (easy, difficulty), and region (Flanders, Brussels, Wallonia).
2. Psychological variables within this domain included self-reported worries about one’s financial situation (low, high), worry about job loss or future employment difficulties (low, high), loneliness (DeJong Gierveld Loneliness Scale with high total score indicate indicates a higher loneliness), and life satisfaction (a scale from 0 to 10 with high total score indicate indicates a higher satisfaction).
3. Other individual-level variables encompassed functional limitations in daily activities due to health problems, and the use of psychotropic medications (sleeping tablets, tranquilizers, anxiolytics, or antidepressants).

**Domain 2: Relationship Factors**: This domain focused on the quality of social support (poor, strong), and the perceived satisfaction with social contacts (satisfying, unsatisfying).

**Domain 3: Societal Factors:** This domain included measures of perceived mental health stigma, assessed using The Stigma-9 Questionnaire (STIG-9) with high total score indicate indicates a higher perceived stigma (18). Additionally, levels of trust in scientific institutions and trust in the national government were categorized as confident or less confident.

Covariates were classified as either time-fixed or time-varying based on their expected stability across waves. Time-fixed variables included demographic characteristics (age, gender, education, region), functional limitations, and psychotropic medication use, which were assumed to remain constant. Time-varying variables, measured in at least two waves to capture change over time, included psychological factors (loneliness, life satisfaction) and social support (Appendix 1).

### Statistical analysis

#### Missing data

Missing values within the dataset arose from two distinct mechanisms: planned missingness (PM) and item nonresponse (INR). Planned missingness was an intentional feature of the study design, as certain measures were administered only at pre-specified survey waves. Such gaps are considered *missing completely at random* (MCAR), as their absence is independent of any observed or unobserved participant characteristics. Item nonresponse is occurring when participants fail to answer specific questions they were administered that could potentially follow MCAR, *missing at random* (MAR), or *missing not at random* (MNAR) patterns. The proportion of missing data due to INR for variables at each wave was examined and is detailed in Appendix 1.

Although PM and INR have different theoretical underpinnings, prior empirical and methodological research suggests that separate imputation strategies for each mechanism seldom yield substantial practical advantages. Consequently, both types of missingness were addressed concurrently within a unified multiple imputation framework (19). This approach maximized the utilization of all available observed data in a single, cohesive procedure. Prior to initiating multiple imputation, for each time-fixed variable, the first available non-missing value for a participant was carried forward to subsequent waves for that same participant if data were missing at those later points.

To accurately reflect the three-level hierarchical structure of the data (repeated observations or waves nested within individuals, who are in turn nested within geographic regions), a fully conditional specification three-level (FCS-3L) imputation strategy was employed (20). Under this paradigm, each incomplete variable was imputed iteratively using a univariate mixed effects model. These models were conditioned on both fully observed covariates and the most recently imputed values of other incomplete variables. All imputation models were fitted using the *ml*.*lmer()* function from the *miceadds* package in R (21). Random intercepts were specified at both the region and individual levels, and additional random slopes for the ‘wave’ variable were included for genuinely time-varying outcomes. To enhance numerical stability and prevent singular model fits, a custom predictor matrix was constructed by retaining only the most strongly correlated covariates for each target variable undergoing imputation. Predictive mean matching was then used to generate realistic imputed values. Ultimately, a total of m = 30 completed datasets were produced, following 10 burn-in iterations. Descriptive sample characteristics (means and standard deviations, or frequencies and percentages) were subsequently pooled from these 30 imputed datasets.

#### Mixed effect latent class analysis

To characterize the temporal evolution of anxiety and depression symptom scores, latent class linear mixed models (LCLMM) were utilized, as implemented in the *lcmm* package for R. Prior to modeling, both anxiety (GAD-7) and depression (PHQ-9) scores were log-transformed (using the formula: log(score + 1)) to handle zeros value and improve the normality of their distributions. Separate *hlme()* models were then fitted for anxiety and depression, testing specifications with one through five latent classes (22). Each model included ‘time’ (wave) as both a fixed effect and a random effect (allowing for random intercepts and random slopes at the individual level to capture individual variability in baseline scores and change over time).

Model selection was guided by a combination of statistical and substantive criteria: minimization of the Bayesian Information Criterion (BIC, and the retention of any identified latent class that comprised at least 5% of the total sample (23). Based on the posterior class-membership probabilities derived from the optimal model, each participant was assigned to their most likely symptom trajectory class. Finally, the resulting latent trajectory groups were cross-tabulated against key demographic variables for descriptive aim.

#### Multilevel mixed effect model

To identify factors associated with anxiety and depression symptom trajectories while accounting for the nested (hierarchical) structure of the data, multilevel mixed effects linear models were applied. Depression and anxiety scores (log-transformed) were modeled as outcomes, with variations attributable to differences within individuals over time (Level 1), between individuals, and between geographic regions (Level 2, though the primary nesting was waves within individuals). Each model incorporated random intercepts for individuals and regions to capture both within- and between-group variability in baseline symptom levels, and a random slope for ‘time’ (wave) to model individual differences in symptom trajectories over the study period.

All covariates were treated as fixed effects and were added to the models sequentially, following the structure of the socio-ecological framework. The analysis commenced with a null model (Model 0), which included only the random-effects structure (random intercepts for individuals and regions, plus a random slope for wave). Subsequently, four nested models were fitted: Model 1 added individual-level demographic and health factors; Model 2 further incorporated individual-level psychological factors; Model 3 introduced relationship factors; and Model 4, the full model, finally included broader societal factors.

Selected time-varying covariates were interacted with ‘time’ (wave) to allow their effects on anxiety and depression symptoms to evolve over the study period. Model fit was compared across this nested sequence of models using the Akaike Information Criterion (AIC), with lower AIC values indicating a better balance between model fit and parsimony. Finally, fixed-effect estimates (coefficients) and their corresponding standard errors from the best-fitting model for both anxiety and depression were pooled across the 30 imputed datasets using Rubin’s rules, as implemented in the *pool()* function of the mice package in R.

## RESULTS

### Demographic characteristics

The cohort is older (mean age ∼56 years), predominantly female, and largely with post‐secondary education. Nearly one‐quarter report financial hardship. Most live as childless or child‐raising couples, while one‐fifth live alone. Geographically, two‐thirds are in Flanders, with smaller representation from Brussels and Wallonia.

The pronounced skew and clustering at low symptom levels indicate that most participants report minimal anxiety or depression, with progressively fewer individuals experiencing moderate to severe symptoms. These distributions justify log transformation in subsequent modeling to meet analytic assumptions and mitigate the influence of extreme values.

### The temporal evolution of anxiety symptoms

The five-class solution demonstrated the best balance between fit and parsimony (lowest BIC), supporting the extraction of five distinct anxiety-symptom trajectories. These classes vary markedly in size—from a small subgroup (8.4%) to the largest class (40.4%) and set the stage for characterizing their longitudinal patterns and associated covariates.

Figure 2 reveals five distinct anxiety‐symptom trajectories over the 2020–2024 period and their uncertainty. Approximately 40 % of participants (Class 3) maintained a steady, mild‐to‐moderate level of anxiety, while a small but concerning 8.4 % (Class 2) began with minimal symptoms that climbed steadily into the moderate‐to‐severe range. A further 20.7 % (Class 1) exhibited persistently minimal symptoms, and 14.0 % (Class 5) experienced chronically severe anxiety with little change. Additionally, 10.4 % of the sample (Class 4) started with high, severe symptoms yet showed pronounced improvement over time.

**Figure 1.**
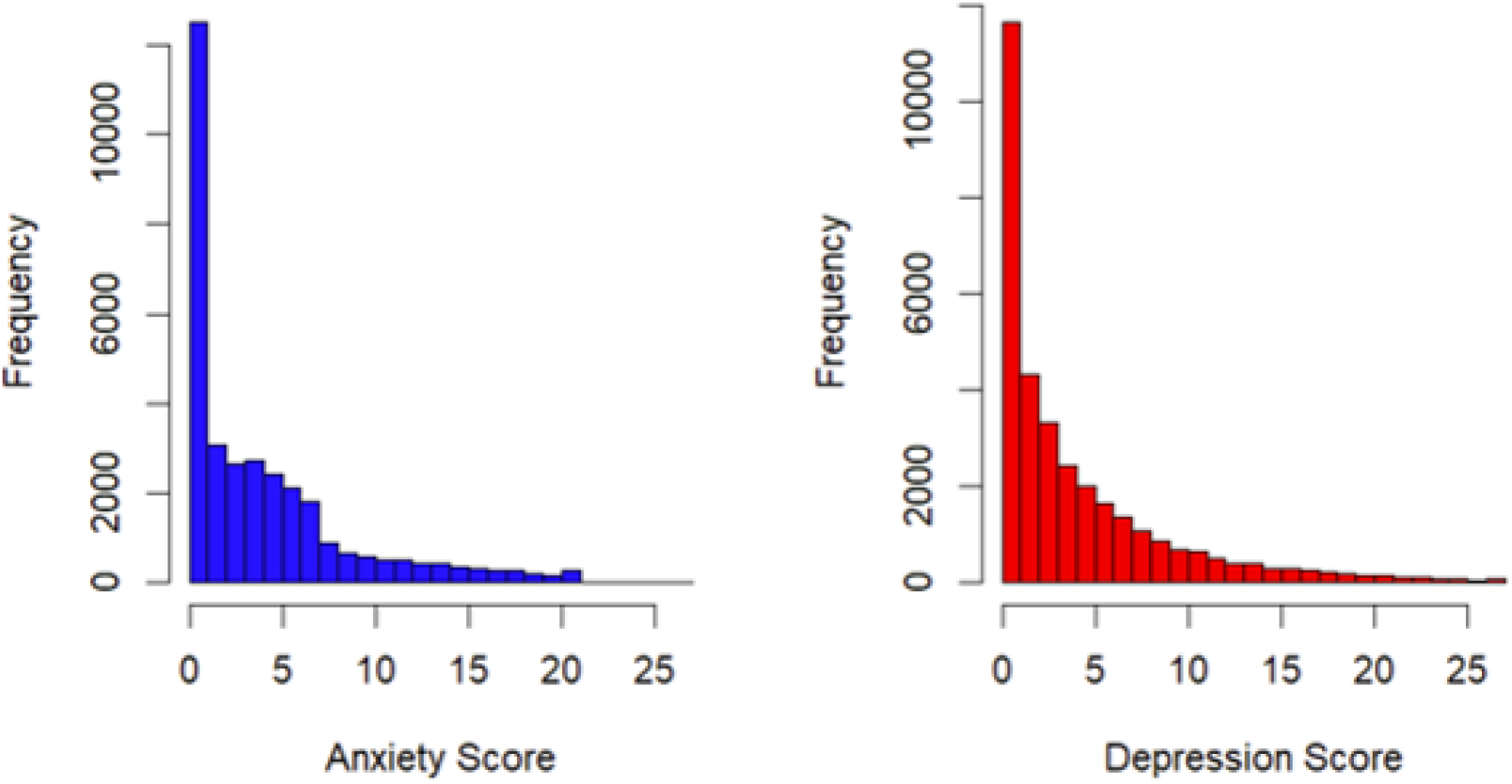
Histogram of Anxiety and Depression score

**Figure 2.**
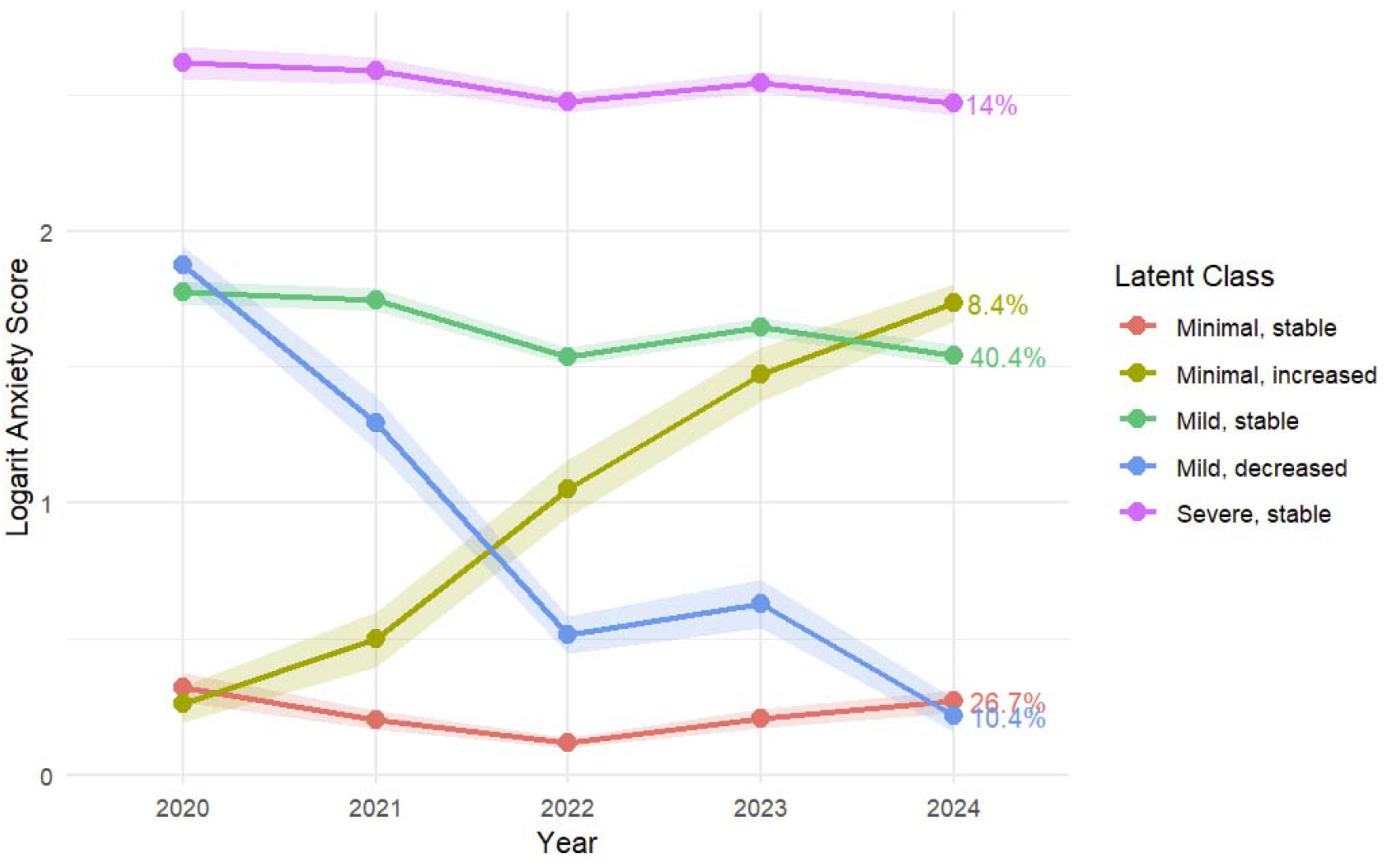
Latent classes of anxiety score

Trajectory membership is strongly stratified by sociodemographic. Stable minimal anxiety (Class 1) is linked to older age, male gender, higher SES, and Flanders residence. In contrast, persistent severe anxiety (Class 5) disproportionately affects younger, female, lower-Perceived income sufficiency individuals in Wallonia, often in non-traditional households.

### The temporal evolution of depression symptom

The four-class solution was selected for its optimal trade-off between model fit and parsimony and for retaining only those trajectories that each comprised at least 5 % of the sample.

Figure 3 depicts four distinct depression‐symptom trajectories from 2020 to 2024 and their uncertainty. Over half of participants (Class 3, 52.3 %) reported persistently mild‐moderate depression with a slight downward trend; about one‐fifth (Class 1, 20.5 %) maintained minimal, stable symptoms throughout; a smaller subgroup (Class 2, 15.9 %) experienced a chronically severe depression with little change; and roughly one in ten (Class 4, 11.3 %) began with minimal symptoms but showed a marked increase into the moderate‐severe range.

**Figure 3.**
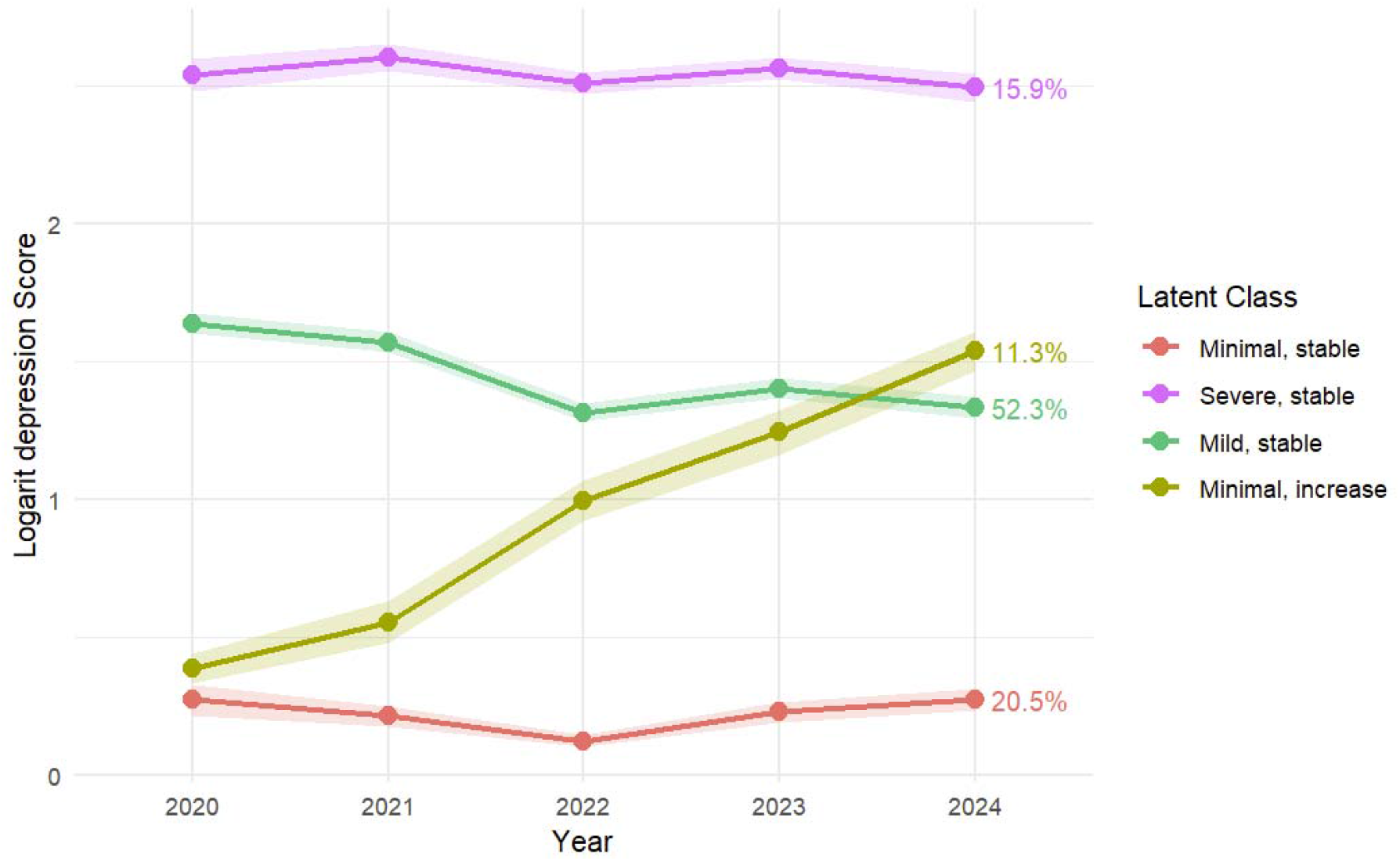
Latent classes of depression score

**Figure 4.**
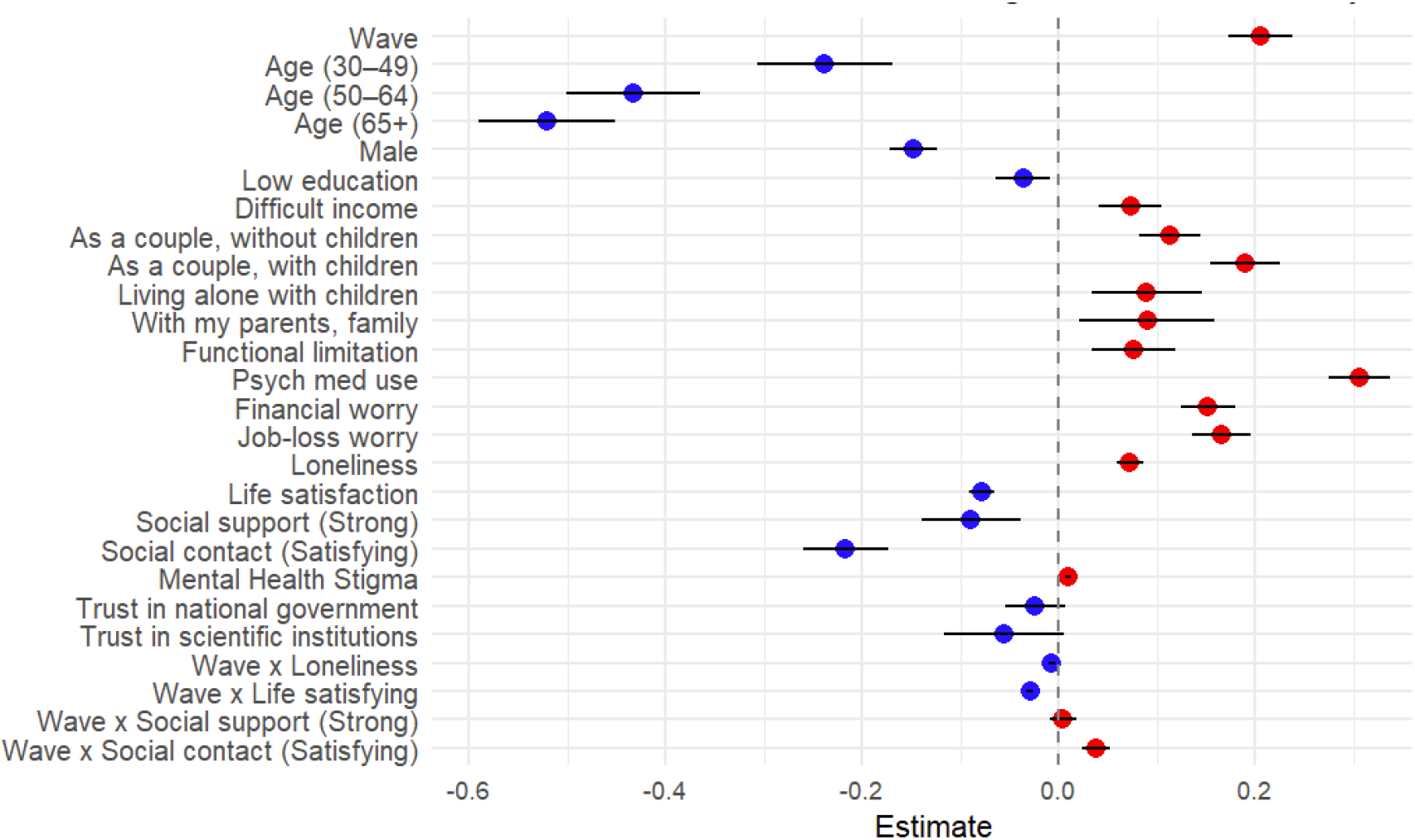
Multilevel linear mixed effect model of Log-transformed anxiety score

**Figure 5.**
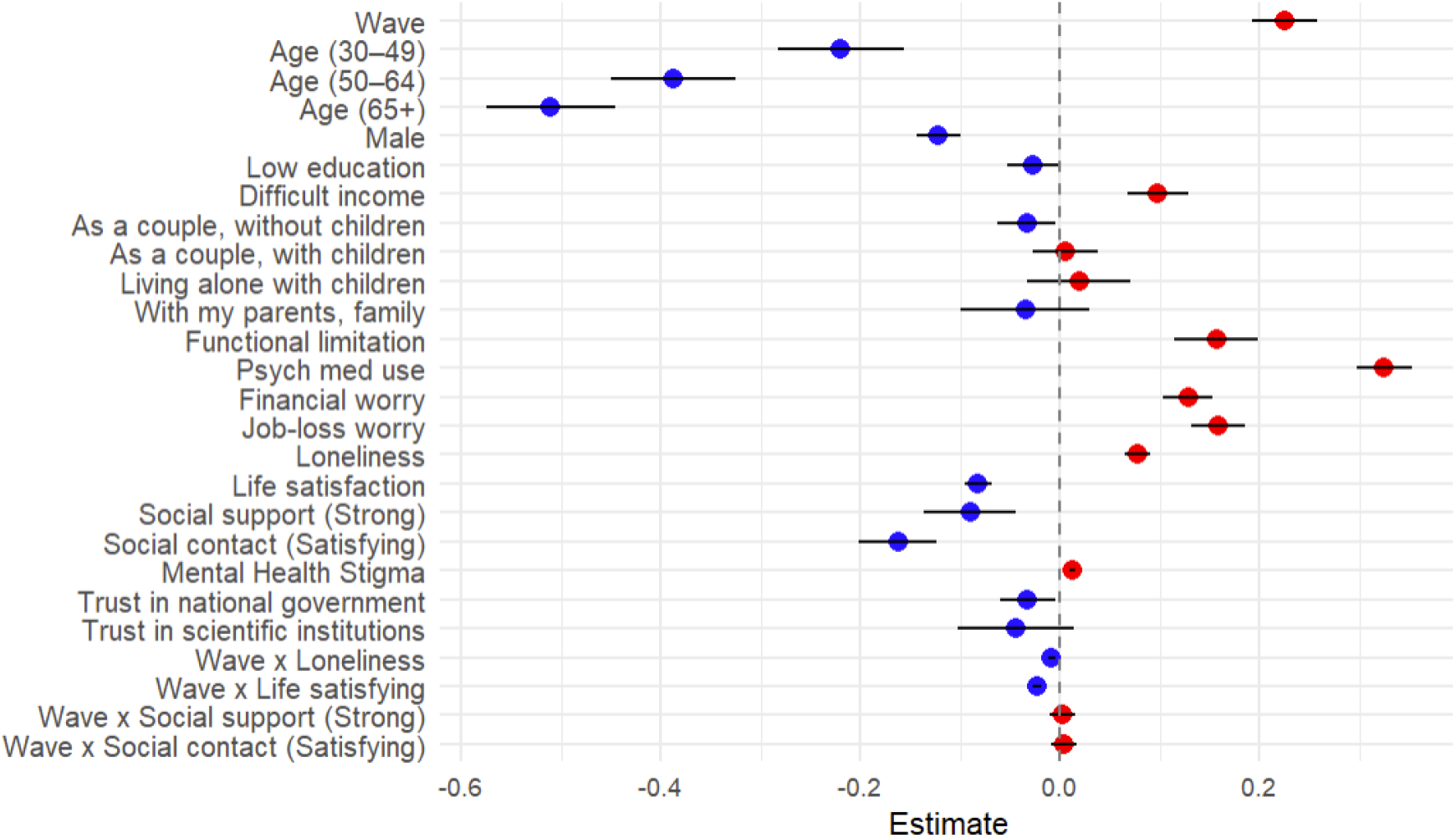
Multilevel linear mixed effect model of Log-transformed depression score

Stable minimal‐symptom individuals are older, male, socioeconomically secure, and concentrated in Flanders. The mild‐decreasing group, despite moderate symptoms, faces notable economic stress and younger age. Chronic severe depression affects a cross‐section of mid‐ to later‐life adults with elevated hardship.

### Multilevel linear mixed effect model

After testing five models, model 4 included all covariates and yielded the lowest AIC (see Appendix 2). This model revealed several significant predictors of anxiety including older age, male gender, higher life satisfaction, strong social support, and satisfying social contact were associated with lower anxiety scores. In contrast, financial worry, job‐loss worry, loneliness, psychotropic medication use, and mental health stigma were linked to higher anxiety. Societal‐level trust in national government and scientific institutions also showed a protective effect. Interaction terms indicated a decline over time in the protective influence of life satisfaction, social support, and social contact.

Older age, male gender, strong social ties, and high life satisfaction were associated with lower depression scores. Conversely, financial and job‐loss worry, loneliness, use of psychotropic medication, and high mental health stigma were associated with higher scores. Trust in national institutions was again protective. Interaction effects revealed a temporal weakening of the benefits of social contact and social support in protecting against depression, suggesting the erosion of these resources over the prolonged crisis period

## DISCUSSION

### Findings and implication

The period from 2020 to 2024 presented unprecedented challenges to global mental health, characterized by the COVID-19 pandemic and multiple intersecting social instabilities. Our study examined both the heterogeneous trajectories of anxiety and depression and the key protective and risk factors across the social-ecological model during this extraordinary period. This discussion brings together those findings to offer a clearer picture of how the Belgian population responded to prolonged adversity.

Our latent class analysis revealed remarkable heterogeneity in mental health responses, identifying five distinct anxiety trajectories and four depression trajectories. For depression, while the majority (52.3%) maintained mild-to-moderate symptoms, smaller but significant groups showed either persistent resilience (20.5%), chronic severity (15.9%), or deterioration (11.3%). Similarly, anxiety manifested in diverse patterns includes steady mild-to-moderate symptoms (40.4%), resilience (26.7%), chronic severity (14.0%), improvement (10.4%), and gradual deterioration (8.4%). This trajectory heterogeneity challenges simplistic pandemic narratives and extends beyond landmark UK study in two critical ways (24). First, our longer observation period (2020-2024) captured important temporal extensions beyond the acute pandemic phase. Second, the proportion of individuals demonstrating resilience in our study (20.5% for depression; 26.7% for anxiety) was substantially lower than previously reported due to the gradual erosion of coping resources over time. People may have been resilient during the early stages of the pandemic but struggled to maintain that resilience as the crisis dragged on and new stressors (24, 25).

We observed a recovery trajectory only in anxiety, with about 10.4% of participants showing symptom improvement. This trend did not appear in the depression data. The absence of a clear recovery for depression is notable, as depression often relates to cumulative losses or past failures, which can require a longer, more multifaceted recovery process than anxiety. However, this finding differs from a systematic review and meta-analysis which showed about 54% of young people with anxiety or depression symptoms recover naturally within a year (26).

When comparing the deterioration trajectories across both conditions, a higher proportion of participants experienced rising symptoms in depression (Class 4, 11.3%) than in anxiety (Class 2, 8.4%). Notably, the increase in anxiety symptoms showed a sharper acceleration during the later years (2022– 2024), whereas the rise in depression followed a more gradual and steadier course throughout the study period. This divergence may reflect distinct psychological response mechanisms that depression may be more reactive to the cumulative impact of sustained loss and disruption, while anxiety may be more closely tied to ongoing uncertainty. These delayed deterioration patterns are particularly concerning, as they are not commonly reported in previous literature. These delayed deterioration patterns are concerning and challenge the common “recovery assumption” in disaster mental health, where distress typically peaks early and then declines (27). Our findings instead support delayed effects, with mental health problems emerging only after prolonged adversity (28). This progressive worsening likely reflects the cumulative psychological toll of the prolonged pandemic and additional societal stressors like economic instability and climate crises. While the demographic profile for this deteriorating trend wasn’t distinct, unmeasured factors such as migration status or physical activities, or even the acute impact of Belgium’s high 2020 COVID-19 mortalities, may have contributed, requiring further research to assess.

Our findings also highlight distinct sociodemographic characteristics associated with severe and mild symptom trajectories. Financial difficulty consistently predicted chronic symptoms, with economically vulnerable individuals overrepresented in severe classes, aligning with the concept of “economic scarring” (29, 30). Gender disparities were also prominent, with females disproportionately represented in mid and severe trajectories, reflecting accumulated disadvantages from structural factors like caregiving burdens and job instability (31-33). While older adults demonstrated resilience, younger adults showed greater anxiety vulnerability, consistent with higher sensitivity to pandemic disruptions. This age-related protection in older adults likely stems from more effective emotion regulation, fewer life disruptions, and broader experience(24, 34, 35).

Our multilevel analysis identified distinct patterns of risk and protection across ecological levels.

#### Individual level factors

In line with the trajectory findings, longitudinal analysis confirmed that older age and male gender were protective factors associated with lower depression and anxiety over time, while financial concerns, particularly subjective financial worry, were consistent risk factors. Compared to earlier studies conducted during the acute phase of the pandemic, our findings suggest that these associations not only persisted but may have strengthened over the extended period, highlighting the long-term mental health impact of both structural disadvantage and perceived economic insecurity (31, 36, 37)

#### Psychological factors

Our findings demonstrate that worry and loneliness emerged as significant risk factors for poorer mental health outcomes. While initial social distancing and disruptions heightened loneliness and isolation, our longitudinal data indicate that associated mental health vulnerabilities persisted amidst prolonged societal disruption (38). Conversely, life satisfaction proved to be a significant protective factor against both depression and anxiety, with its beneficial effect increasing over time. This suggests that interventions should not only promote initial life satisfaction but also help individuals sustain and deepen it, as its protective benefits are cumulative (39, 40).

#### Relationship level factors

In both anxiety and depression models, strong perceived social support and satisfying social contact were significantly associated with lower symptom levels. These results align with extensive research showing that supportive social relationships buffer individuals from the psychological impacts of stress and adversity (41). However, the observed interaction effects indicate that the strength of these protective associations declined over time. One possible explanation is supporting fatigue or erosion, a phenomenon where the psychological benefits of social resources weaken during prolonged periods of shared stress (30, 42). Another plausible explanation is that individuals may have developed internal coping mechanisms and naturally recovered independently, reducing their reliance on social factors for emotional regulation (43). These findings highlight the need for time-sensitive mental health interventions that provide early social support while fostering long-term individual resilience.

#### Societal level factors

Mental health stigma, measured using the Stigma-9 Questionnaire, was positively associated with higher levels of both anxiety and depression. This suggests that individuals with stronger stigmatizing beliefs toward mental illness may be less likely to acknowledge symptoms or seek help, leading to worse psychological outcomes (44, 45). Trust in national government and scientific institutes is associated with lower scores, highlighting the protective role of institutional confidence. When individuals believe that their government is acting competently and in their best interest, it may reduce uncertainty and anxiety during prolonged disruption. Trust in science may support more adaptive responses to health messaging, reduce susceptibility to misinformation, and foster a sense of control (46).

### Study strengths and limitations

This study possesses several notable strengths. Firstly, its longitudinal design, with five data collection points spanning over five years, provides a rare and valuable opportunity to track the dynamic evolution of mental health symptoms from the acute phase of the pandemic into the post-pandemic era. This allows for a more nuanced understanding than cross-sectional studies. Secondly, the large, national sample size, derived from Sciensano’s established health surveys, enhances the robustness of the findings for understanding mental health trajectories and associated factors within this cohort. However, it is crucial to consider the socio-demographic composition, as women comprise 61.80% of the study sample (Table 2), a higher proportion than in the general adult Belgian population (approximately 50.8%). While this study did not aim to report population prevalence, this demographic imbalance suggests that the observed trajectories and associations might not be directly transferable or applicable in the same proportions to all subgroups within the broader population (47). Thirdly, our classification of variables as time-fixed or time-varying provides important insights into the stability of influences across ecological levels. Demographic characteristics created relatively stable patterns of differential vulnerability, while psychological factors and social resources showed greater fluctuation in their effects over time. This temporal distinction suggests what might be termed “ecological adaptation which different levels of the social ecology adjust to prolonged disruption at varying rate.

**Table 2.** Characteristics of study participants (n=10,063)

| Variable | Frequency (n) | Percentage (%) |
| --- | --- | --- |
| Age (mean ± SD) | ± 55.8 (13.7) | |
| Age group |  |  |
| 18 - 29 | 423 | 4.20 |
| 30 - 49 | 2690 | 26.73 |
| 50 - 64 | 3829 | 38.05 |
| 65+ | 3121 | 31.01 |
| Gender (Female) | 6219 | 61.80 |
| High education<br>(Higher secondary school diplomas) | 7431 | 73.84 |
| **Perceived income sufficiency** (Difficult) | 2603 | 25.87 |
| Household type |  |  |
| Living alone | 2149 | 21.36 |
| As a couple, without child(ren) | 4107 | 40.81 |
| As a couple, with child(ren) | 2702 | 26.85 |
| Living alone with children | 549 | 5.46 |
| With my parent(s), family, friends or acquaintances | 441 | 4.38 |
| Other | 115 | 1.14 |
| Region |  |  |
| Flanders | 6630 | 65.88 |
| Brussels | 958 | 9.52 |
| Wallonia | 2475 | 24.60 |

**Table 3.** Model fit statistics for latent class analysis of anxiety symptom trajectories.

| Model | G | loglik | npm | BIC | class1 | class2 | class3 | class4 | class5 |
| --- | --- | --- | --- | --- | --- | --- | --- | --- | --- |
|  |  |  |  |  | % | % | % | % | % |
| 2-Class | 2 | -38011.7 | 10 | 76115.65 | 42.35 | 57.65 |  |  |  |
| 3-Class | 3 | -37565 | 14 | 75258.97 | 23.97 | 50.62 | 25.41 |  |  |
| 4-Class | 4 | -37399.8 | 18 | 74965.5 | 24.13 | 37.16 | 27.19 | 11.53 |  |
| **5-Class** | **5** | **-37336.4** | **22** | **74875.55** | **26.68** | **8.45** | **40.45** | **10.43** | **13.99** |

**Table 4.** Demographic and socioeconomic characteristics by anxiety latent class (n=10,063)

| Class 1 | Class 2 | Class 3 | Class 4 | Class 5 |
| --- | --- | --- | --- | --- |
| n = 2,685 | n = 850 | n = 4,070 | n = 1,050 | n = 1,408 |

|  |  | Minimal,<br>stable | Minimal,<br>increased | Mild, stable | Mild,<br>decreased | Severe,<br>stable |
| --- | --- | --- | --- | --- | --- | --- |
| Age group |  |  |  |  |  |  |
|  | 18-29 | 33 (1.2) | 27 (3.2) | 213 (5.2) | 35 (3.3) | 115 (8.2) |
|  | 30 - 49 | 363 (14) | 198 (23) | 1,309 (32) | 279 (27) | 541 (38) |
|  | 50 - 64 | 1,011 (38) | 317 (37) | 1,510 (37) | 447 (43) | 544 (39) |
|  | 65+ | 1,278 (48) | 308 (36) | 1,038 (26) | 289 (28) | 208 (15) |
| Gender |  |  |  |  |  |  |
|  | Female | 1,292 (48) | 517 (61) | 2,732 (67) | 701 (67) | 977 (69) |
|  | Male | 1,393 (52) | 333 (39) | 1,338 (33) | 349 (33) | 431 (31) |
| Education |  |  |  |  |  |  |
|  | High educated | 1,932 (72) | 597 (70) | 3,172 (78) | 769 (73) | 961 (68) |
|  | Low educated | 753 (28) | 253 (30) | 898 (22) | 281 (27) | 447 (32) |
| Perceived income sufficiency |  |  |  |  |  |  |
|  | Easily | 2,295 (85) | 629 (74) | 2,956 (73) | 832 (79) | 748 (53) |
|  | With difficulty | 390 (15) | 221 (26) | 1,114 (27) | 218 (21) | 660 (47) |
| Household type |  |  |  |  |  |  |
|  | Living alone | 601 (22) | 208 (24) | 836 (21) | 218 (21) | 286 (20) |
|  | As a couple, without<br>child(ren) | 1,399 (52) | 364 (43) | 1,480 (36) | 431 (41) | 433 (31) |
|  | As a couple, with<br>child(ren) | 501 (19) | 197 (23) | 1,252 (31) | 308 (29) | 444 (32) |
|  | Living alone with children | 98 (3.6) | 41 (4.8) | 253 (6.2) | 41 (3.9) | 116 (8.2) |
| With my parent(s), family,<br>friends or acquaintances |  | 61 (2.3) | 30 (3.5) | 202 (5.0) | 39 (3.7) | 109 (7.7) |
|  | Other | 25 (0.9) | 10 (1.2) | 47 (1.2) | 13 (1.2) | 20 (1.4) |
| Region |  |  |  |  |  |  |
|  | Flanders | 1,986 (74) | 571 (67) | 2,515 (62) | 734 (70) | 832 (59) |
|  | Brussels | 216 (8.0) | 74 (8.7) | 438 (11) | 81 (7.7) | 133 (9.4) |
|  | Wallonia | 483 (18) | 205 (24) | 1,117 (27) | 235 (22) | 443 (31) |

**Table 5.** Model fit statistics for latent class analysis of depression symptom trajectories.

| Model | G | loglik | npm | BIC | class1 | class2 | class3 | class4 | class5 |
| --- | --- | --- | --- | --- | --- | --- | --- | --- | --- |
|  |  |  |  |  | % | % | % | % | % |
| 2-Class | 2 | -35368.6 | 10 | 70829.29 | 19.30 | 80.70 |  |  |  |
| 3-Class | 3 | -35193.8 | 14 | 70516.57 | 19.16 | 63.88 | 16.96 |  |  |
| **4-Class** | **4** | **-35130** | **18** | **70425.89** | **20.50** | **15.87** | **52.32** | **11.31** |  |
| 5-Class | 5 | -35104 | 22 | 70410.83 | 21.14 | 1.48 | 9.57 | 47.67 | 20.15 |

**Table 6.**
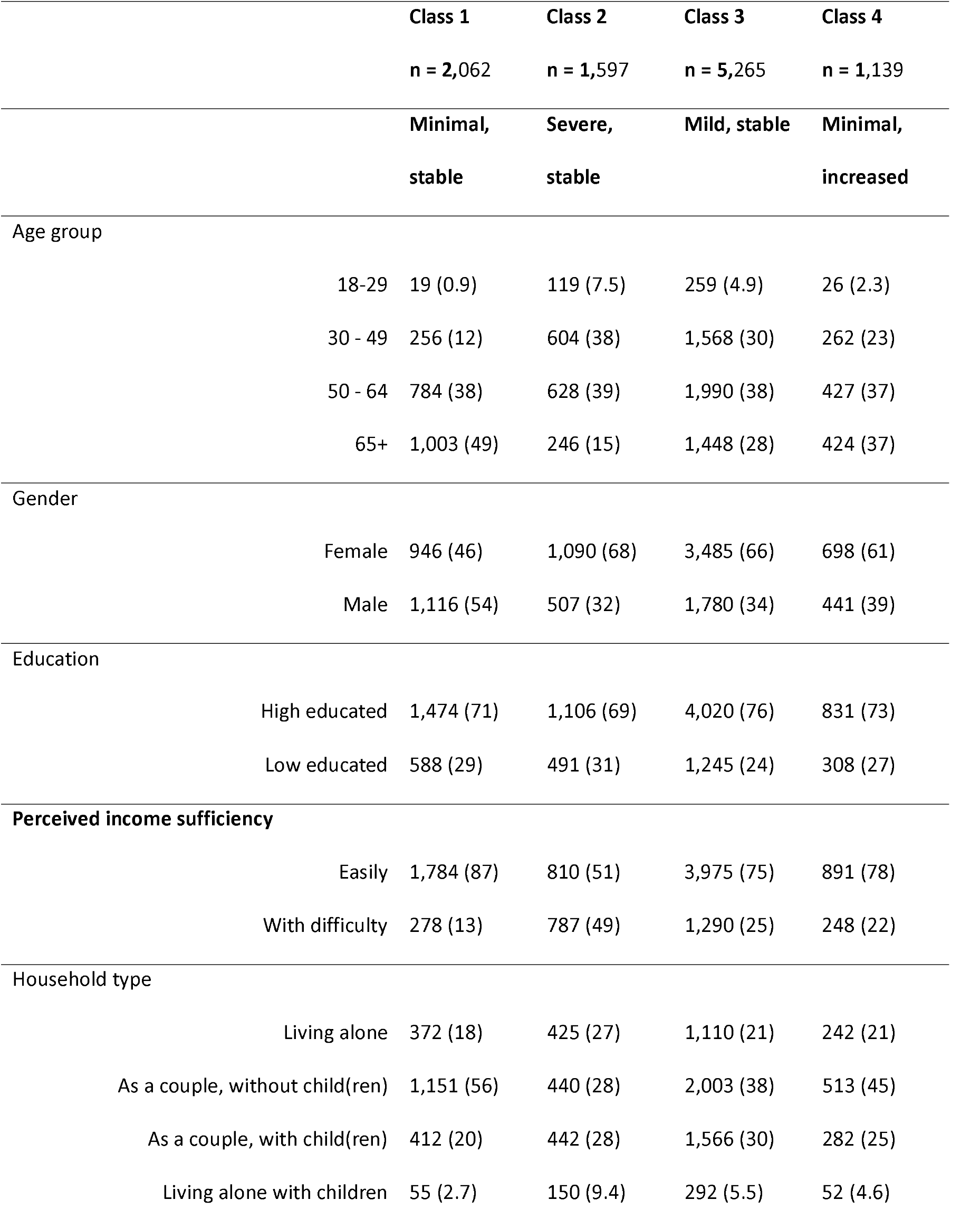
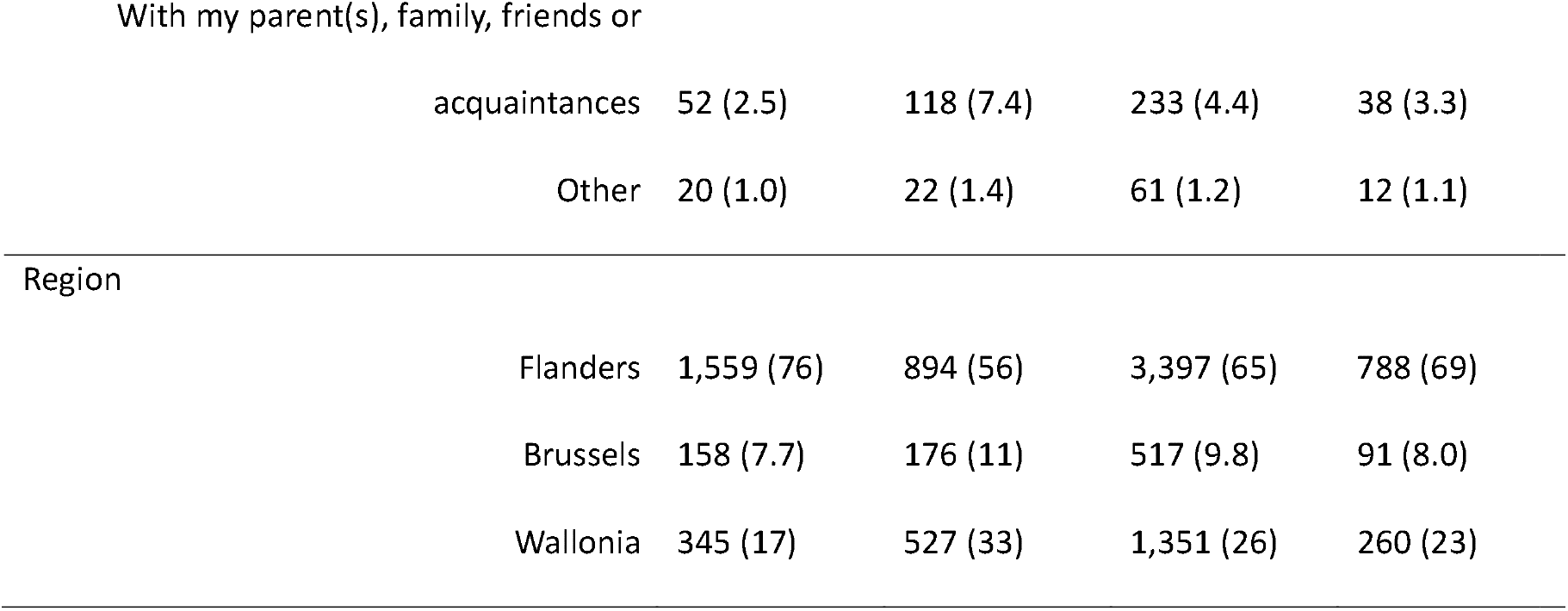
Demographic and socioeconomic characteristics by depression latent class(n=10,063)

Despite its strengths, this study is subject to certain limitations. Firstly, the reliance on self-reported measures for anxiety and depression, while common in large-scale surveys, may be subject to reporting biases such as social desirability and recall bias. Clinical diagnostic interviews were not feasible in this survey context. Secondly, while the BELHEALTH cohort aimed for longitudinal follow-up, the initial COVID-19 Health Surveys were based on convenience samples. This could introduce selection bias, potentially limiting the representativeness of the initial cohort, although efforts were made to make it as representative as possible. Thirdly, attrition over the multiple waves of data collection is a common challenge in longitudinal studies. While statistically addressed through imputation and mixed models, it could still affect the results if attrition was systematically related to unobserved characteristics.

### Conclusions and recommendation

In conclusion, this study provides a comprehensive picture of mental health responses in Belgium from 2020 to 2024, revealing diverse trajectories of anxiety and depression.. The persistence of psychological distress among significant subgroups, particularly women, younger adults, and those facing financial hardship, underscores the need for targeted and sustained public mental health strategies. Interventions should address not only individual psychological risk factors like loneliness and low life satisfaction but also broader structural determinants such as economic insecurity, stigma, and institutional trust. Given the changing of protective effects of social and psychological resources over time, future mental health policies must be dynamic, combining early outreach with long-term resilience-building efforts across all levels of the social-ecological model.

## Data Availability

The datasets used and/or analysed during the current study are available via Sciensano at on reasonable request.

## List of abbreviations

COVID-19: Coronavirus Disease 2019
GAD-7: Generalized Anxiety Disorder 7-item scale
PHQ-9: Patient Health Questionnaire 9-item scale
AIC: Akaike Information Criterion
WHO: World Health Organization
STIG-9: The Stigma-9 Questionnaire
PM: Planned missingness
INR: Item nonresponse
MCAR: Missing Completely At Random
MAR: Missing At Random
MNAR: Missing Not At Random
FCS-3L: Fully Conditional Specification Three-Level
LCLMM: Latent Class Linear Mixed Models
BIC: Bayesian Information Criterion

## DECLARATIONS

## Ethics approval and consent to participate

Informed consent was obtained from all subjects involved in the study. The COVID-19 Health Surveys and the BELHEALTH study were approved by the ethics committee of the Ghent university hospital (Commissie voor Medische Ethiek), B.U.N. B6702020000031. The study was conducted in accordance with the Declaration of Helsinki.

## Consent for publication

N/A

## Clinical trial number

not applicable. This study is a longitudinal observational study and not an interventional clinical trial

## Authors’ contributions

S.D., C.D., and G.V. contributed to the conceptualization of the study and the development of the methodology. S.D. and C.D. were responsible for data collection, while C.D. managed the data. T.B. performed the data analysis, formal statistical analyses and prepared the original draft of the manuscript. S.D., C.D., and G.V. reviewed and edited the manuscript critically. S.D., C.D., and G.V. also supervised the overall project. All authors have read and agreed to the published version of the manuscript.

## Acknowledgements

The authors would like to express their sincere gratitude to Epidemiology and Public Health, Sciensano, for their invaluable contribution in providing the data for this study. We are also deeply thankful to the Department of General Practice and Population Health, University of Antwerp for their insightful comments and constructive feedback throughout this research. We also thank the participants who responded to the surveys.

## Supplementary Information

## APPENDIX 1

### Missing data percentage

| Variable | Type | Wave 1 | Wave 2 | Wave 3 | Wave 4 | Wave 5 |
| --- | --- | --- | --- | --- | --- | --- |
| ID | Structure | 0.00 | 0.00 | 0.00 | 0.00 | 0.00 |
| wave | Structure | 0.00 | 0.00 | 0.00 | 0.00 | 0.00 |
| Region | Structure | 0.00 | 0.00 | 0.00 | 0.10 | 0.03 |
| Age | Time-fixed | 0.00 | 0.00 | 0.00 | 0.32 | 0.00 |
| Gender | Time-fixed | 0.00 | 0.00 | 0.00 | 0.16 | 0.00 |
| Education | Time-fixed | 0.00 | 0.00 | 0.00 | 0.14 | 0.08 |
| Household type | Time-fixed | 0.00 | 0.00 | 0.00 | 0.49 | 0.77 |
| Anxiety score | Time-fixed | 14.20 | 8.32 | 5.76 | 0.79 | 1.10 |
| Depression score | Time-fixed | 15.83 | 8.48 | 5.79 | 0.84 | 1.16 |
| Functional limitation | Time-fixed | 0.00 | - | - | - | - |
| Used any sleeping tablets,<br>tranquilizers or anxiolytic | Time-fixed | - | - | - | 0.78 | 1.07 |
| Used any antidepressants | Time-fixed | - | - | - | 0.54 | 1.52 |
| Financial worry | Time-fixed | - | - | 0.00 | 0.99 | - |
| Job-loss worry | Time-fixed | - | - | 0.00 | 26.41 | - |
| Loneliness | Time-varying | - | 0.00 | 0.00 | - | 1.95 |
| Life satisfaction | Time-varying | - | 4.21 | 2.27 | 0.25 | 0.29 |
| Social support | Time-varying | 0.00 | 0.00 | 0.00 | 2.73 | 4.87 |
| Social contact | Time-varying | 9.73 | 8.23 | 6.80 | 0.97 | 2.25 |
| Mental Health Stigma | Time-fixed | - | - | - | - | 5.81 |
| Trust in national government | Time-fixed | 0.00 | - | - | - | - |
| Trust in scientific institutions | Time-fixed | 0.00 | - | - | - | - |

### Little’s MCAR test

Little's MCAR test
| statistic | df | p.value | missing. patterns |
| --- | --- | --- | --- |
| <dbl> | <dbl> | <dbl> | <int> |
| 80991.31 | 24550 | 0 | 1183 |

### Histogram of missing-data proportions

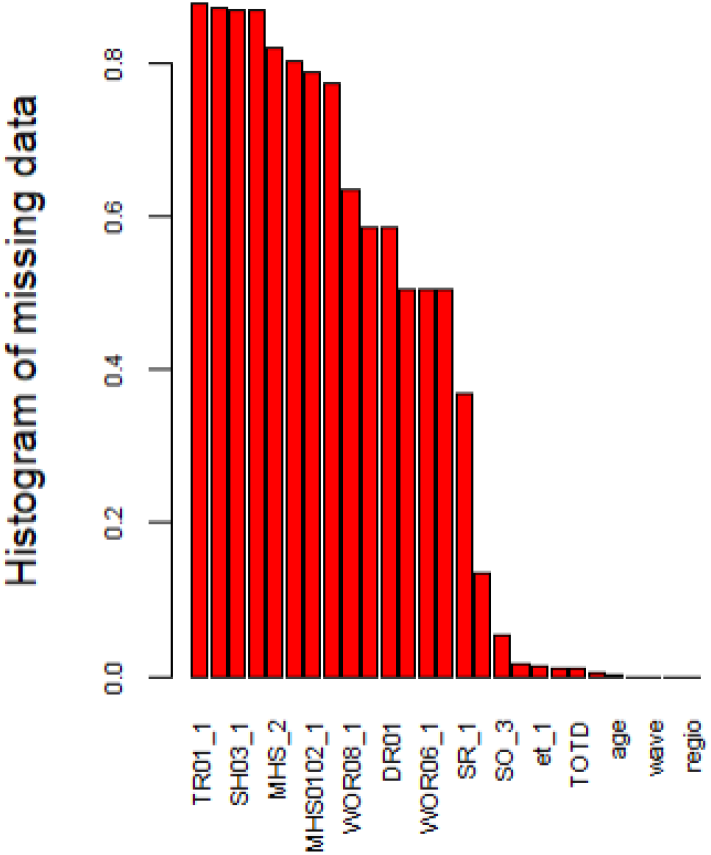

## APPENDIX 2

### Multilevel linear mixed effect model of Log-transformed anxiety score

| Variable | Null Model<br>b (SE) | Model 1 | Model 2 | Model 3 | Model 4 |
| --- | --- | --- | --- | --- | --- |
| Fixed effects |  |  |  |  |  |
| *Constant* | 1.33 (0.3) | 1.65 (0.05) | 1.8 (0.07) | 1.89 (0.07) | 1.82 (0.08) |
| *Wave* | -0.23 (0.01) | -0.03 (0) | 0.22 (0.02) | 0.2 (0.02) | 0.21 (0.02) |
| *Individual factors* |  |  |  |  |  |
| Age |  |  |  |  |  |
| 30 - 49 |  | -0.26 (0.04) | -0.24 (0.04) | -0.23 (0.03) | -0.24 (0.04) |
| 50 - 64 |  | -0.53 (0.04) | -0.43 (0.03) | -0.42 (0.03) | -0.43 (0.03) |
| 65+ |  | -0.75 (0.04) | -0.53 (0.04) | -0.51 (0.04) | -0.52 (0.04) |
| Gender (Male) |  | -0.14 (0.02) | -0.14 (0.01) | -0.15 (0.01) | -0.15 (0.01) |
| Education (high) |  | -0.01 (0.02) | -0.03 (0.01) | -0.05 (0.01) | -0.04 (0.01) |
| Perceived income<br>sufficiency difficult |  | 0.35 (0.02) | 0.08 (0.02) | 0.01 (0.02) | 0.07 (0.02) |
| Household type |  |  |  |  |  |
| Couple, no child(ren) |  | 0.03 (0.02) | 0.12 (0.02) | 0.12 (0.02) | 0.11 (0.02) |
| Couple, with child(ren) |  | 0.11 (0.02) | 0.19 (0.02) | 0.19 (0.02) | 0.19 (0.02) |
| Living alone with<br>children |  | 0.1 (0.04) | 0.09 (0.03) | 0.09 (0.03) | 0.09 (0.03) |
| Living with<br>parent(s)/family |  | 0.1 (0.04) | 0.09 (0.04) | 0.08 (0.03) | 0.09 (0.04) |
| Functional limitation |  | 0.18 (0.03) | 0.08 (0.02) | 0.07 (0.02) | 0.08 (0.02) |
| Psych med use |  | 0.48 (0.02) | 0.31 (0.02) | 0.3 (0.02) | 0.31 (0.02) |
| *Psychological factors* |  |  |  |  |  |
| Financial worry |  |  | 0.16 (0.01) | 0.14 (0.01) | 0.15 (0.01) |
| Job-loss worry |  |  | 0.18 (0.02) | 0.12 (0.02) | 0.17 (0.02) |
| Loneliness |  |  | 0.09 (0.01) | 0.07 (0.01) | 0.07 (0.01) |
| Life satisfaction |  |  | -0.09 (0.01) | -0.08 (0.01) | -0.08 (0.01) |
| Loneliness * Wave |  |  | -0.01 (0) | -0.01 (0) | -0.01 (0) |
| Life satisfaction * Wave |  |  | -0.03 (0) | -0.03 (0) | -0.03 (0) |
| *Relationship factors* |  |  |  |  |  |
| Social support (Strong) |  |  |  | -0.09 (0.03) | -0.09 (0.03) |
| Social contact (Satisfying) |  |  |  | -0.22 (0.02) | -0.22 (0.02) |
| wave × Social support<br>(Strong) |  |  |  | 0 (0.01) | 0 (0.01) |
| wave × Social contact<br>(Satisfying) |  |  |  | 0.04 (0.01) | 0.04 (0.01) |
| *Societal factors* |  |  |  |  |  |
| Mental Health Stigma |  |  |  |  | 0.01 (0) |
| Trust in national<br>government |  |  |  |  | -0.02 (0.02) |
| Trust in scientific<br>institutions |  |  |  |  | -0.06 (0.03) |
| Random effect |  |  |  |  |  |
| Random Intercept<br>(Subject) | 13.29 (3.70) | 11.26 (3.36) | 5.60 (2.37) | 5.67 (2.38) | 5.66 (2.38) |
| Random Intercept<br>(Region) | 0.244 (0.49) | 0.11 (0.32) | 0.04 (0.2) | 0.04 (0.19) | 0.02 (0.15) |
| Random Slope (Wave) | 0.18 (0.12) | 0.10 (0.08) | 0.15 (0.11) | 0.12 (0.09) | 0.14 (0.10) |
| AIC | 82314 | 79520 | 71142 | 70725 | 70609 |

### Multilevel linear mixed effect model of Log-transformed depression score

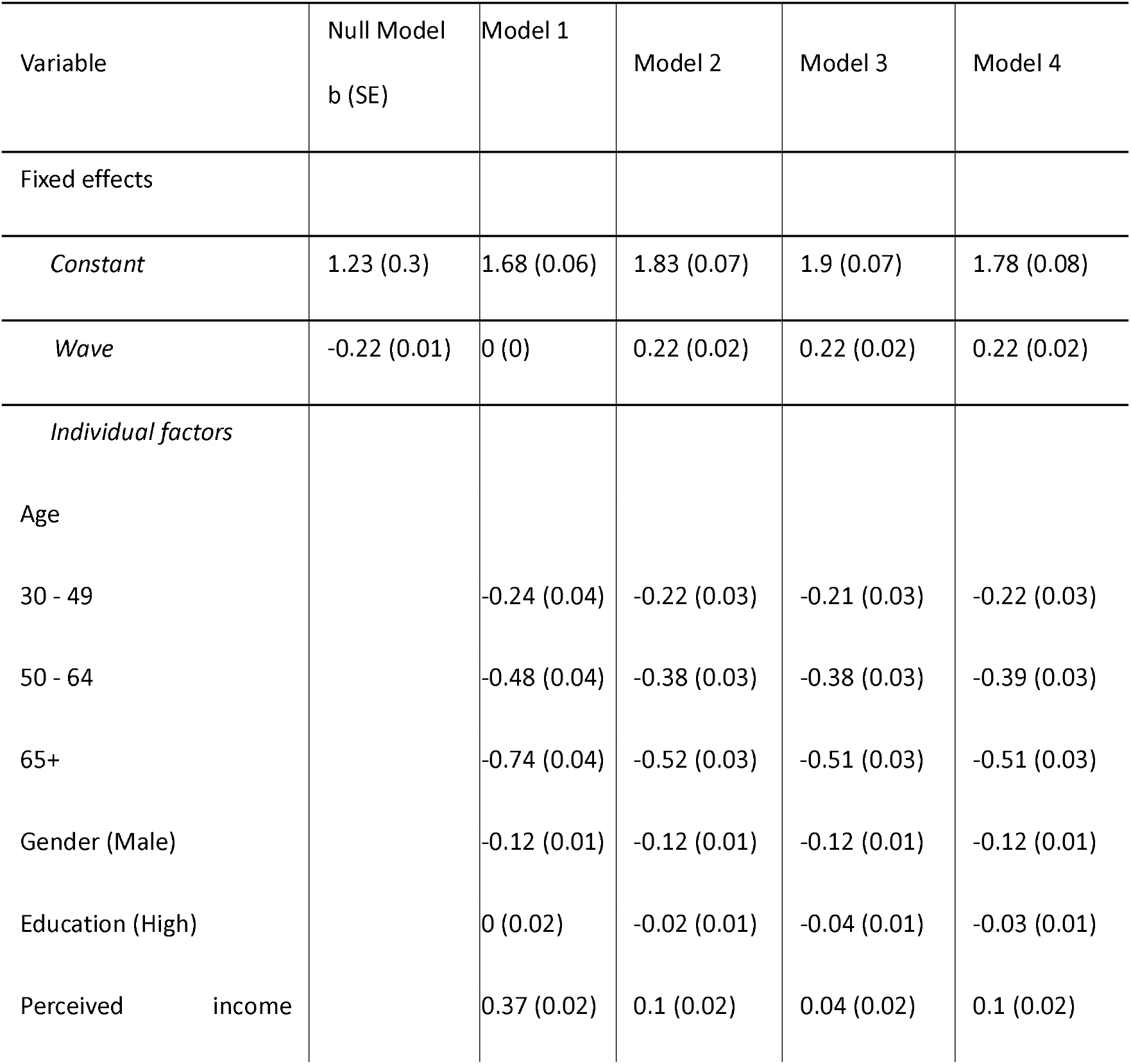
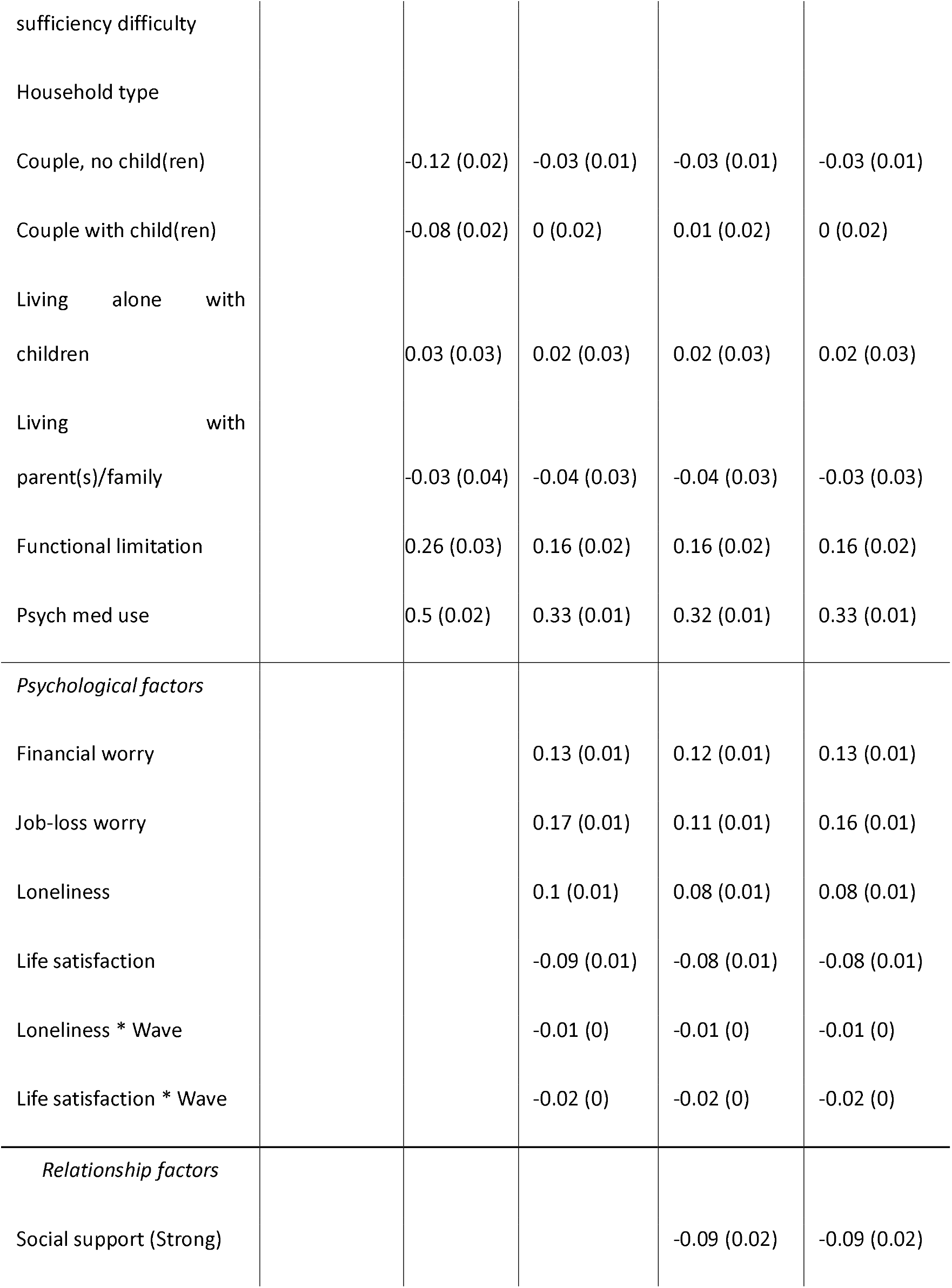
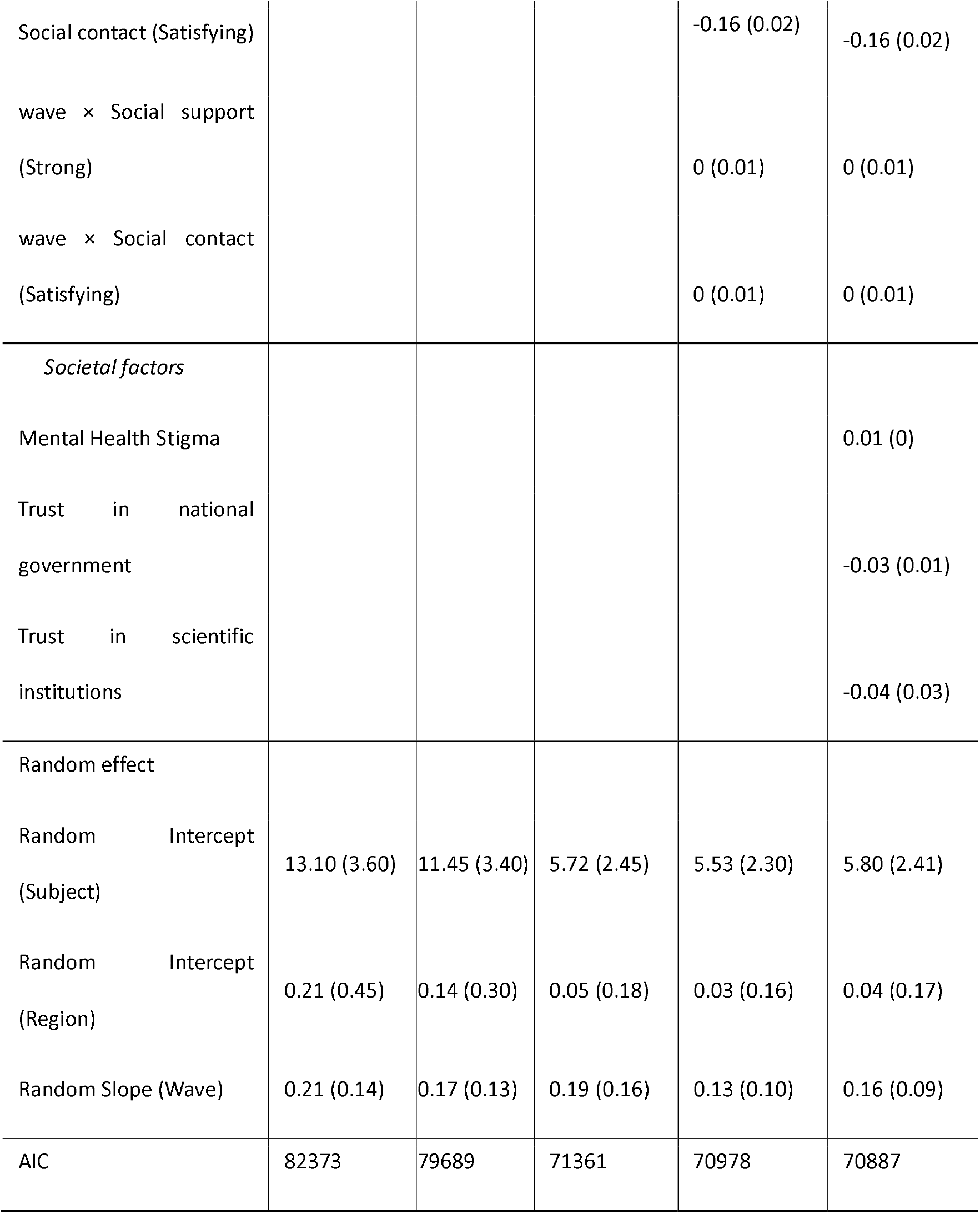

